# Utility of the ADAS-Cog as a Cognitive Screening Tool in Older Adults with Epilepsy: A Multicenter Cohort Study

**DOI:** 10.64898/2026.05.27.26354210

**Authors:** Jessica Kania, Ifrah Zawar, Anny Reyes, Victoria Williams, Rani Sarkis, Vineet Punia, McKenna Williams, Lisa Ferguson, Kayela Arrotta, Robyn M Busch, Jana Jones, Bruce P Hermann, Carrie R McDonald

**Affiliations:** Department of Neurology, University of Wisconsin School of Medicine and Public Health, Madison WI, USA; Department of Neurology, University of Virginia School of Medicine, Charlottesville VA, USA; Epilepsy Center and Department of Neurology, Neurological Institute, Cleveland Clinic, Cleveland OH, USA; Department of Medicine, University of Wisconsin School of Medicine and Public Health, Madison WI, USA; Department of Neurology, Harvard Medical School, Brigham and Women’s Hospital, Boston MA, USA; Departments of Psychiatry and Radiation Medicine and Applied Sciences, University of California-San Diego, San Diego CA, USA

**Keywords:** epilepsy, ADAS-Cog, neuropsychology, screening

## Abstract

**Objective:** Cognitive impairment is common among older adults with epilepsy, although efficient screening tools suitable for routine use are lacking. Here we examine, for the first time, the utility of the Alzheimer’s Disease Assessment Scale-Cognitive Subscale (ADAS-Cog) as a screening tool to identify cognitive impairment in older adults with epilepsy.

**Methods:** Participants included 83 adults (ages ≥ 55) with epilepsy from the Brain, Aging, and Cognition in Epilepsy (BrACE) study and 83 age-, sex-, and education-matched cognitively healthy controls from the Alzheimer’s Disease Neuroimaging Initiative (ADNI-3). All completed the ADAS-Cog and a comprehensive neuropsychological battery to identify cognitive phenotypes (intact vs impaired). Performance on individual ADAS-Cog items and the total score was assessed, and diagnostic efficiency statistics were determined.

**Results:** Epilepsy participants (mean age=66.4 years) performed significantly worse across the ADAS-Cog total score and 8 of the 13 individual test items compared to controls. The largest effect sizes were observed on verbal learning and memory tasks, particularly word recall (*d*=0.87) and delayed word recall (*d*=1.06). An ADAS-Cog total score of ≥15 yielded optimal diagnostic efficiency (67.5% accuracy, 68.8% sensitivity, 66.7% specificity) for identifying cognitive impairment.

**Significance:** The ADAS-Cog is sensitive to detecting cognitive impairment in older adults with epilepsy and may represent a scalable screening option in this population. Additional comparative studies in older epilepsy populations are needed to determine the sensitivity of this measure to longitudinal change, cross-cultural applicability, and availability across languages.

**Plain language summary:** Cognitive decline is common among older adults with epilepsy, although sufficient evidence supporting the use of screening tools to identify cognitive impairment in this population is lacking. The ADAS-Cog may be a useful screening option in epilepsy research and clinical care, although additional studies are needed to compare it with other cognitive screening tests and to confirm its applicability for clinical care and across cultures and healthcare settings.

**Key Points:**

□ ADAS-Cog, a tool widely used in Alzheimer’s disease research, was used to detect cognitive impairment in older adults with epilepsy versus healthy controls.
□ Largest deficits were in verbal learning and delayed recall, aligning with epilepsy-related memory vulnerability.
□ ADAS-Cog was compared with the gold standard, the International Classification of Cognitive Disorders in Epilepsy (IC-CoDE). An ADAS-Cog-13 cutoff ≥15 showed optimal accuracy for IC-CoDE–defined cognitive impairment.
□ ADAS-Cog performance reliably distinguished IC-CoDE-impaired from intact participants within the epilepsy cohort.
□ These findings support ADAS-Cog as a cognitive screening tool in aging epilepsy populations to identify neurodegenerative vulnerability.

## Introduction

Epilepsy and Alzheimer’s disease (AD) are highly prevalent neurological disorders with a bidirectional relationship (1,2). Older adults represent the fastest-growing segment of epilepsy prevalence worldwide, with epilepsy increasingly recognized as a major comorbidity that carries a significantly increased risk of developing Alzheimer’s disease and related dementias (ADRD) (3–6). In AD, seizures were traditionally viewed as a later consequence of the disease (3,4), but it is now known that seizures can occur early in the AD course (7–9). These findings have led to the proposal of an epileptiform phenotype of AD characterized by hippocampal and mesial temporal lobe hyperexcitability, detectable EEG abnormalities, subtle seizures, and accelerated cognitive change (10). On the other hand, older adults with a history of either chronic active epilepsy (11) or remitted childhood-onset epilepsy (36,37) show a higher rate of AD biomarker positivity, suggesting epilepsy may also predispose older individuals to developing neurodegenerative pathology.

In older adults, cognitive deficits due to epilepsy in the presence of co-existing dementia pathologies are clinically consequential, being associated with higher mortality, accelerated cognitive decline, and worse functional outcomes (11,12). The extent of cognitive deficits in epilepsy is known to influence medication adherence, seizure safety, functional independence, and quality-of-life (13–16). Despite the substantial impact of cognitive impairment on long-term prognosis, cognitive screening remains inconsistently implemented in the routine care of older adults with epilepsy

Given the aging of the world population and the increased incidence of epilepsy in older adults, the ILAE (International League Against Epilepsy) has recommended cognitive screening for those with late-onset incident epilepsy and aging persons with established epilepsy (17). Efforts to address this recommendation are in their infancy. To date, there has been only one validity study of the Montreal Cognitive Assessment (MoCA) in aging persons with epilepsy (PwE) (18) and no validity studies using the Alzheimer’s Disease Assessment Scale–Cognitive Subscale (ADAS-Cog) either independently or in comparison to other available cognitive screening measures, including the MoCA (18). Given the lack of consensus regarding the applicability of these measures to aging PwE, it is critical to demonstrate the sensitivity and validity of these measures in aging epilepsy populations.

The ADAS-Cog is a standardized cognitive measure extensively used in ADRD research and clinical trials. (12–14). Because epilepsy is routinely specified as an exclusionary criterion for AD intervention trials, the sensitivity of the ADAS-Cog to epilepsy and epilepsy outcomes is unknown. The ADAS-Cog may be particularly relevant to assessing cognition in aging epilepsy populations, given that memory deficits, especially in episodic learning and delayed recall, are frequently observed and may reflect a convergence of epilepsy-related network disruption, accelerated brain aging, and comorbid neurodegenerative pathology (19,20). Additionally, the ADAS-Cog’s widespread use in AD research raises the possibility that it could provide a translational bridge between epilepsy and dementia frameworks, particularly as disease-modifying therapies increase the importance of early identification. With a growing interest in early intervention trials that target pathology prior to the overt clinical dementia syndrome and/or preclinical conditions based on the presence of biomarkers (21), identifying cognitive outcome measures to detect potential treatment effects in milder stages of disease (including mild cognitive impairment[MCI]) is of relevance, particularly for research and clinical trials.

We assessed the utility of the 13-item version of the ADAS-Cog (ADAS-Cog-13) in older adults with focal epilepsy from the multicenter Brain, Aging, and Cognition in Epilepsy (BrACE) cohort. In this first systematic examination of the ADAS-Cog-13 in epilepsy, we (1) compared total and item-level ADAS-Cog-13 performance in aging individuals with focal epilepsy to propensity-matched cognitively healthy older adults from the Alzheimer’s Disease Neuroimaging Initiative-3 (ADNI-3) to assess sensitivity to epilepsy-related cognitive differences; (2) evaluated the concurrent validity of ADAS-Cog-13 performance by benchmarking abnormality against an epilepsy-specific external criterion, the International Classification of Cognitive Disorders in Epilepsy (IC-CoDE) (22); and (3) conducted diagnostic efficiency analyses to identify candidate ADAS-Cog-13 cutoff scores that optimize detection of IC-CoDE–defined cognitive impairment.

## Methods

### Participants

Participants were included from the BrACE study, a multi-site investigation involving the University of California-San Diego (UCSD), Cleveland Clinic (CC), and University of Wisconsin-Madison (UWM) (5,23). BrACE is a prospective, longitudinal study investigating cognition, brain aging, and biomarkers in older adults with focal epilepsy. Inclusion criteria were age ≥ 55 years, English proficiency, diagnosis of focal epilepsy by a board-certified neurologist with expertise in epileptology in accordance with the criteria defined by the ILAE, and no history of prior epilepsy neurosurgery (e.g., resection, neuromodulation device, laser ablation). Exclusion criteria were a diagnosis of a primary neurological disorder (other than epilepsy), a history of significant psychiatric disorder (e.g., psychosis) requiring hospitalization, or a diagnosis of dementia. The project was approved by the UCSD Institutional Review Board and other respective sites, and written informed consent was acquired from all participants.

### Control participants

All controls were cognitively healthy older adults from the ADNI-3 database. PwE were 1:1 matched to ADNI controls using propensity scores based on age, sex, and education. To ensure all BrACE participants received a control match, a 5-year age range and a 2-year education range were used. Independent t-tests and Pearson’s Chi-Square analyses confirmed that no significant differences existed between the two groups in age, education, and sex (all p’s >0.05).

### Clinical and sociodemographic measures

In BrACE, sociodemographic variables (e.g., sex, race, ethnicity, and education) were collected from participant self-reports and health records. Epilepsy clinical history was obtained from a comprehensive clinical interview at the baseline visit and review of available medical records.

### Neuropsychological measures

All BrACE recruitment sites administered the ADNI-3 test battery and additional measures, which consisted of the ADAS-Cog-13, the MoCA, and comprehensive neuropsychological measures of memory, executive function, language, and processing speed. **Supplementary Table 1** presents the administered neuropsychological tests, descriptions of the measures, and the norms used in analyses.

The ADAS-Cog-13 is a standardized measure adapted from the original ADAS-Cog, which includes 11 tasks, including word recall and recognition, naming, following commands, constructional praxis, ideational praxis, orientation, task reminders, and language skills, including spontaneous spoken language ability (intelligibility and clarity of speech in an open conversation), comprehension, and word finding (24). The 13-item version adds a number cancellation task and a delayed free recall trial for the word list (32). Administration of the ADAS-Cog-13 takes between 25 and 35 minutes. Scores range from 0 to 85 and are based on the number of errors, with higher scores indicating greater impairment. The ADAS-Cog is a widely used tool that measures the severity of cognitive impairment in patients with AD and assesses treatment-related changes in mild-to-moderate AD, and is commonly used in AD treatment trials (29,31).

The MoCA is a screening measure used to classify cognitive impairment (25). It is a widely recognized tool sensitive to cognitive impairment that assesses multiple domains (executive function, naming, attention, language, abstraction, memory, and orientation). We have previously validated its utility in the BrACE cohort (23). The MoCA total score was corrected for education by adding one point if the participant had 12 or fewer years of education (25,26).

### IC-CoDE Classification

The IC-CoDE is a consensus-based taxonomy that standardizes the diagnosis of cognitive dysfunction in epilepsy and is used to determine cognitive phenotypes from neuropsychological assessment (22). Based on the administered test battery, four of the five domains were represented, including memory, language, processing speed/attention, and executive function. Phenotypes within the IC-CoDE were classified as *intact* (no impairment across all domains*), single-domain impairment* (one cognitive domain impaired), *bi-domain* (two cognitive domains impaired), and *generalized impairment* (three or more cognitive domains impaired). Impairment in a single domain was defined by abnormality (≥1 standard deviation from the normative means used) across two or more tests within the domain (38). Since the IC-CoDE was used here as a tool to determine the ADAS-Cog’s utility for detecting impairment, the single, bi-domain, and generalized domains were combined into one group for this study and labeled as *IC-CoDE impaired,* while individuals with no impairment across domains were labeled as *IC-CoDE intact*.

### Statistical analyses

Analyses were conducted using IBM SPSS version 30, and the IC-CoDE portal(27) was used to derive cognitive phenotypes. Independent t-tests assessed ADAS-Cog-13 performance in BrACE participants versus ADNI controls for individual tests and overall performance. Cohen’s d effect sizes are reported for significant differences in ADAS-Cog scores between groups. Given the lack of literature defining the optimal ADAS-Cog-13 threshold for impairment in epilepsy, we examined cutoff scores for MCI.

Analyses of sensitivity, specificity, positive predictive value (PPV), negative predictive value (NPV), Kappa, and Youden Index were conducted to determine the accuracy between IC-CoDE and ADAS-Cog cutoff impairment classifications. Receiver operating characteristic (ROC) curves were calculated to determine the area under the curve (AUC) for the potential ADAS-Cog-13 cutoffs. Independent sample t-tests, Analysis of Variance (ANOVA), Pearson’s Chi-square, and Fisher’s exact tests were conducted to analyze ADAS-Cog-13 data to IC-CoDE impairment, as well as sociodemographic and clinical measures. Significant results were reported at the .05 level.

## Results

### Participants

The final sample included 83 older adults with focal epilepsy from the BrACE cohort, for which their demographic and clinical epilepsy characteristics are provided in **Table 1**. Most participants were classified as having active epilepsy (65.8%), defined as more than one seizure a year or having had at least one seizure within the past year.

**Table 1.** Survey of Demographics for BrACE Participants.

|  |  |
| --- | --- |
| **Group (N = 83)** |  |
| **Years of Age** M (SD) | 66.37 (6.84) |
| **Years of Education** M (SD) | 14.88 (2.61) |
| **Epilepsy Onset Age** (years, M (SD)) | 43.24 (20.64) |
| **Epilepsy Duration** (years, M (SD)) | 24.23 (20.32) |
| **ADI National %** M (SD) | 48.18 (26.28) |
| **Number ASMs** M (SD) | 1.67 (0.91) |
| **Sex** |  |
| **Female (%)** | 47 (56.6) |
| **Male (%)** | 36 (43.4) |
| **Race** |  |
| **White (%)** | 73 (88.0) |
| **Black (%)** | 4 (4.8) |
| **Asian (%)** | 1 (1.2) |
| **Native American/Alaskan (%)** | 1 (1.2) |
| **Multiracial (%)** | 2 (2.4) |
| **Ethnicity** |  |
| **Non-Hispanic (%)** | 78 (94.0) |
| **Hispanic (%)** | 5 (6.0) |
| **Epilepsy Onset** |  |
| **Typical (%)** | 58 (70.7) |
| **Late (%)** | 24 (29.3) |
| **Epilepsy Status** |  |
| **Active (%)** | 53 (63.9) |
| **Controlled (%)** | 26 (31.3) |
| **Localization** |  |
| **Frontal (%)** | 4 (4.8) |
| **Temporal (%)** | 41 (49.4) |
| **Frontotemporal (%)** | 21 (25.3) |
| **Unknown (%)** | 17 (20.5) |

### ADNI Controls and BrACE Participants

This epilepsy cohort had higher (worse) ADAS-Cog-13 total scores (M = 14.41, SD = 4.95) compared to the age-, sex-, and education-matched ADNI controls (M = 8.98, SD = 4.31); t(164) = −7.228, p < .001 (*d* = −1.12). Eight of the thirteen test items administered in the ADAS-Cog showed significantly worse performance in the epilepsy cohort compared to controls: word recall (t(164) = −5.576, p < .001, *d* = −0.87); delayed word recall (t(164) = −6.795, p < .001, *d* = −1.06); word recognition (t(164) = −2.284, p = .012, *d* = −0.36); object naming (t(164) = −2.292, p = .012, *d* = −0.36); task reminders (t(164) = −2.862, p = .003, *d* = −0.44); comprehension of spoken language (t(164) = −2.011, p = .024, *d* = −0.31); word finding difficulty (t(164) = −3.597, p < .001, *d* = −0.56); number cancellation task (t(164) = −3.133, p = .001, *d* = − 0.49). Commands, constructional praxis, ideational praxis, orientation, and language were comparable across the groups.

### IC-CoDE Impairment and the ADAS-Cog-13

**Supplementary Table 2** presents the sociodemographic and epilepsy clinical characteristics of the *IC-CoDE impaired* and *IC-CoDE intact* groups. The number of anti-seizure medications (ASMs) was significantly different between groups, with *IC-CoDE impaired* taking a higher average number of ASMs than *IC-CoDE intact,* t(81) = −1.733, p = .044.

Of the 83 BrACE participants, 32 (38.6%) were classified as *IC-CoDE impaired*. The cognitive phenotype with the highest rate of impairment was in the memory domain, with 19 (22.9%) of the 32 impaired individuals, followed by language (18.1% impairment), attention/processing speed (10.8% impairment), and executive function (4.8% impairment).

The *IC-CoDE impaired* group had significantly higher (more abnormal) ADAS-Cog-13 total scores than the *IC-CoDE intact* group, t(81) = −3.172, p = .001, *d* = −0.72 (**Figure 1**). Investigation of IC-CoDE phenotypes with the ADAS-Cog-13 total scores showed significant differences between groups, F(2, 80) = 5.337, p = .007. Post Hoc comparisons with Bonferroni correction indicated that the mean score for the *intact* group (M = 12.93, SD = 4.95) was significantly different from both the *single-domain* group (M = 15.85, SD = 4.46) and the *bi-domain/generalized* group (M = 17.28, SD = 4.93). However, the *single-domain and bi-domain/generalized groups did not differ significantly* from one another. Examining the *IC-CoDE memory-impaired* group, those with memory impairment had more abnormal ADAS-Cog-13 total scores, t(81) = −3.444, p < .001, *d* = −0.90, and ADAS-Cog delayed word recall scores, t(81) = −4.593, p < .001, *d* = −1.20, than those in the *IC-CoDE memory-intact* group.

**Figure 1.**
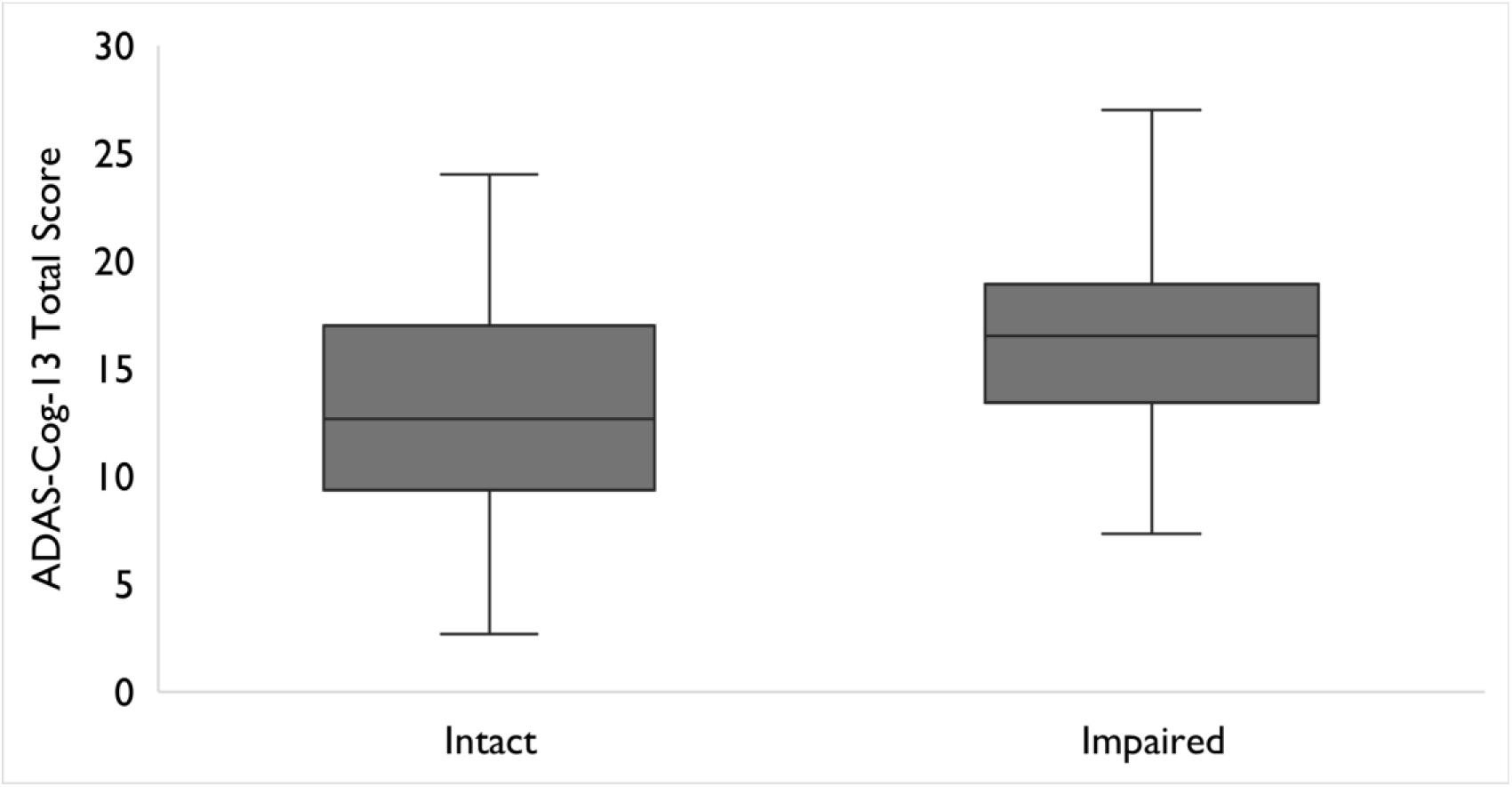
ADAS-Cog-13 (Alzheimer’s Disease Assessment Scale-Cognition 13-item) Total Score vs. IC-CoDE (International Classification of Cognitive Disorders in Epilepsy) Impairment Status

### Detection of IC-CoDE Impairment Using ADAS-Cog-13 Cutoffs

ADAS-Cog-13 total scores ranging from 14 to 18 were examined as cutoffs for cognitive impairment in epilepsy based on their sensitivity, specificity, PPV, NPV, Kappa, and Youden Index to the IC-CoDE classification(**Table 2**). In the sample, 50.6% of participants had an ADAS-Cog-13 total score ≥14.5, 47.0% of participants had ≥15.0, and 39.8% had ≥15.5. ROC plots and their corresponding AUC can be seen in **Figure 2**. ADAS-Cog-13 cutoff scores ranging from 14 to 16 yielded sufficient AUC scores, with ≥15.0 yielding the highest AUC (0.677). The total score cutoff of ≥15.0 yielded the highest accuracy of impairment classification to the IC-CoDE (67.5%), the highest Kappa statistic (0.340), and the best balance between sensitivity/specificity and PPV/NPV across all tested possible cutoffs.

**Figure 2.**
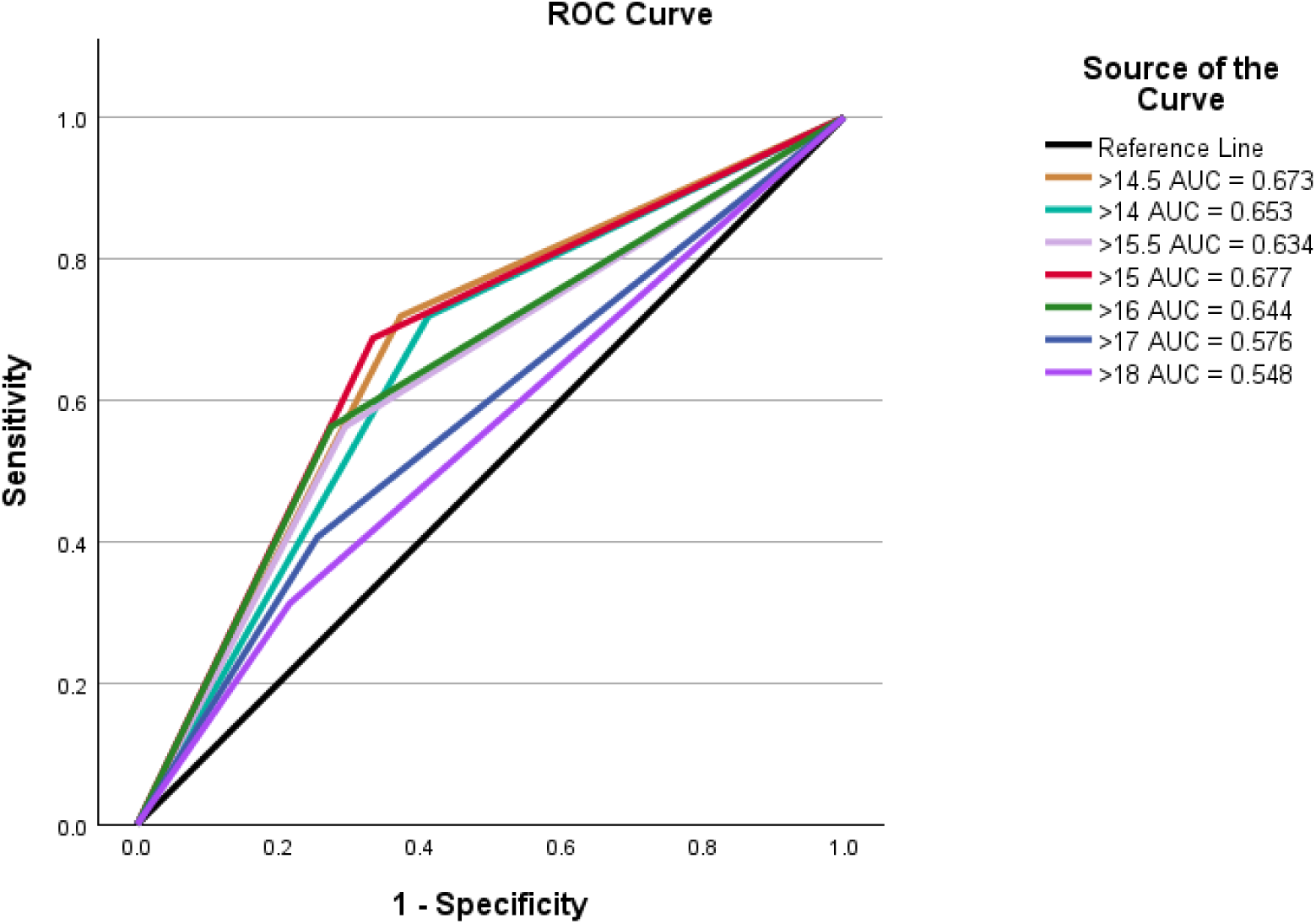
Receiver Operating Characteristic (ROC) Plots for ADAS-Cog-13 (Alzheimer’s Disease Assessment Scale-Cognition 13-item) Cutoffs with Area Under the Curve (AUC)

**Table 2.** Sensitivity and Specificity Analysis of the ADAS-Cog-13 Potential Cutoffs to IC-CoDE Classification.

| **Cut-off Value** | **% of sample impaired** | **Accuracy** | **Sensitivity** | **Specificity** | **PPV** | **NPV** | **Kappa** | **Youden Index** |
| --- | --- | --- | --- | --- | --- | --- | --- | --- |
| **18.0** | 25.3% | 60.2% | 31.2% | 78.4% | 47.6% | 64.5% | 0.103 | 0.097 |
| **17.0** | 31.3% | 61.4% | 40.6% | 74.5% | 50.0% | 66.7% | 0.157 | 0.151 |
| **16.0** | 38.6% | 66.3% | 56.3% | 72.5% | 56.3% | 72.5% | 0.288* | 0.288 |
| **15.5** | 39.8% | 65.1% | 56.3% | 70.6% | 54.5% | 72.0% | 0.267* | 0.268 |
| **15.0** | **47.0%** | **67.5%** | **68.8%** | **66.7%** | **56.4%** | **77.3%** | **0.340\*** | **0.354** |
| **14.5** | 50.6% | 66.3% | 71.9% | 62.7% | 54.8% | 78.0% | 0.327* | 0.346 |
| **14.0** | 53.0% | 63.9% | 71.9% | 58.8% | 52.3% | 76.9% | 0.287* | 0.307 |

Note: \* indicates a significant Kappa of $p < .05$ .

### ADAS-Cog-13 Performance and Relation to Sociodemographic and Clinical Factors

**Supplementary Table 3** shows independent sample t-tests and Spearman Rho correlations between the ADAS-Cog-13 total scores and sociodemographic and clinical factors. Male epilepsy participants had higher (more abnormal) ADAS-Cog-13 total scores compared to females, t(81) = −1.986, p = .025, *d* = − 0.44. MoCA total scores had a significant negative correlation to ADAS-Cog-13 total scores, *r_s_*(81) = −.40, p < .001. Other sociodemographic factors, such as area deprivation index (ADI) national percentile, education, and age, were not significant. Epilepsy clinical factors, such as age of onset, duration, seizure activity, and number of ASMs, were not significant predictors of ADAS-Cog-13 total performance.

### ADAS Cog-13 compared to MoCA

**Table 3** provides a direct comparison of the two screening tests regarding their diagnostic efficiency in this cohort. MoCA outperformed the ADAS-Cog to a modest but detectable degree across the metrics presented.

**Table 3.**
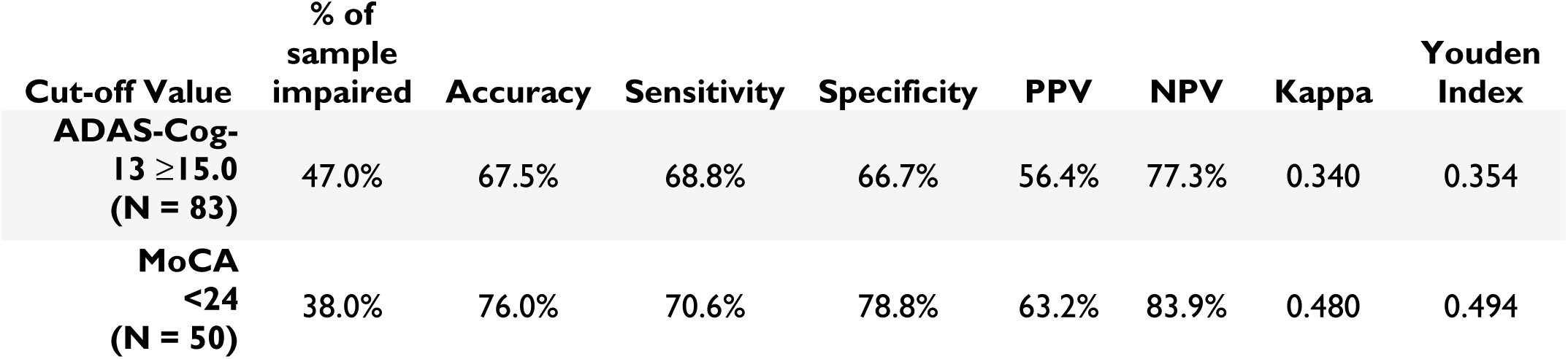
Comparison of Classification Accuracy to the IC-CoDE between the ADAS-Cog-13 and the MoCA.

| Cut-off Value | % of sample impaired | Accuracy | Sensitivity | Specificity | PPV | NPV | Kappa | Youden Index |
| --- | --- | --- | --- | --- | --- | --- | --- | --- |
| **ADAS-Cog-13 $\geq 15.0$**<br>**(N = 83)** | 47.0% | 67.5% | 68.8% | 66.7% | 56.4% | 77.3% | 0.340 | 0.354 |
| **MoCA <24**<br>**(N = 50)** | 38.0% | 76.0% | 70.6% | 78.8% | 63.2% | 83.9% | 0.480 | 0.494 |

## Discussion

In this multicenter study, we demonstrate that the ADAS-Cog-13 shows acceptable sensitivity and specificity in detecting cognitive impairment in older adults with focal epilepsy. Compared with age-, sex-, and education-matched cognitively healthy controls from ADNI-3, epilepsy participants performed significantly worse on the ADAS-Cog-13 total score and on the majority of individual subtests, with the largest effect sizes observed in verbal learning and delayed recall. Within the epilepsy cohort, higher ADAS-Cog-13 scores (i.e., worse performance) reliably distinguished individuals classified as cognitively impaired by the IC-CoDE, particularly those with memory impairment. An ADAS-Cog-13 cutoff score of ≥15 yielded the most favorable balance of sensitivity, specificity, and overall diagnostic accuracy for identifying cognitive impairment in this population. Together, these findings support the ADAS-Cog-13 as a potential screening measure for cognitive impairment in older adults with epilepsy.

A key finding is that the direct group comparisons of ADAS-Cog-13 performance demonstrated significantly worse total scores in epilepsy participants compared to controls, supporting the overall sensitivity of the instrument in this population, but with meaningful variability across discrete items. No significant group differences were observed on items assessing commands, constructional praxis, ideational praxis, orientation, or spontaneous spoken language. In contrast, significant impairments in the epilepsy group were evident across all memory-related tasks (word recall, delayed recall, word recognition, task reminders), as well as subtests assessing nominal language (object naming, word-finding difficulty), comprehension of spoken language, and number cancellation. As the participant population is predominantly composed of temporal lobe pathology, the items showing significant impairment support the region of dysfunction. The non-significant items lean towards the assessment of parietal function. Effect sizes further underscore the domain-specific sensitivity of the ADAS-Cog-13. Word recall and delayed word recall demonstrated large effect sizes (>0.8), word-finding difficulty showed a medium effect size (>0.5), and remaining affected subtests exhibited small but meaningful effects (>0.2). These findings suggest that the ADAS-Cog is not only sensitive to memory and language vulnerabilities preferentially impacted by AD pathology, but is also sensitive to cognitive functioning patterns common in epilepsy.

The prominent deficits observed in verbal learning and delayed recall are consistent with prior literature demonstrating that memory dysfunction is a common cognitive phenotype in older adults with focal epilepsies, even among those without a diagnosis of dementia. Prior work has also shown that compared to amnestic MCI, older PwE exhibit prominent impairments in language (naming and fluency) (28) as well as processing speed and executive function (29). Multiple studies have shown that episodic memory impairment in epilepsy may reflect a convergence of epilepsy-related network dysfunction, cumulative seizure burden, medication effects, and age-related or neurodegenerative pathology (19,20). The large effect sizes observed for ADAS-Cog word recall and delayed recall tasks suggest that these subtests are particularly sensitive to some of the cognitive vulnerabilities most relevant to aging epilepsy populations. Importantly, these memory subtests align closely with those outlined in the IC-CoDE framework, reinforcing the construct validity of the ADAS-Cog when applied to epilepsy.

Classification of epilepsy participants using IC-CoDE revealed substantial heterogeneity, with 61% classified as cognitively intact, 24% with single-domain impairment, 11% with bi-domain impairment, and 4% with generalized impairment. Memory was the most frequently affected domain, followed by attention/processing speed and language. This heterogeneity raises important questions regarding the efficient identification of individuals at greatest cognitive risk and the role of screening tools in guiding referral for comprehensive neuropsychological assessment. To address this, participants were dichotomized into IC-CoDE intact versus any IC-CoDE impaired. Epilepsy characteristics, including age of onset, epilepsy duration, seizure activity, age, and education, were largely unrelated to impairment status, with the exception of a modest association with greater ASM burden. These findings align with prior work suggesting that cognitive impairment in older adults with epilepsy is not solely explained by seizure-related factors, but reflects a multifactorial process involving brain aging and comorbid pathology (20)(34).

From a clinical utility perspective, identifying an optimal ADAS-Cog-13 cut-point is critical. A major strength of this study is the direct benchmarking of ADAS-Cog-13 performance against IC-CoDE classifications derived from comprehensive neuropsychological testing. By anchoring the ADAS-Cog-13 to an epilepsy-specific standard, this work provides meaningful estimates of diagnostic efficiency rather than relying solely on group differences. Further, an ADAS-Cog-13 cutoff score of ≥15 demonstrated the most favourable balance of sensitivity, specificity, predictive values, and overall classification accuracy. Although the AUC was modest (0.677), this performance is consistent with expectations for a brief screening measure applied to a heterogeneous clinical population and is comparable to commonly used cognitive screeners in epilepsy (30).

In direct comparison with prior MoCA data from a subset of this cohort (20), supplemented here to match our ADAS-Cog-13 cohort (**Table 3**), the MoCA demonstrated higher overall classification accuracy and AUC. This finding does not diminish the potential utility of the ADAS-Cog in aging PwE as the two instruments differ substantially in domain weighting, intended use, comprehensiveness, and translational relevance (30,31). The ADAS-Cog-13 places greater emphasis on episodic memory and delayed recall, domains that showed the largest effect sizes in the present study and that are central to both epilepsy-related cognitive vulnerability and neurodegenerative risk. Unlike the MoCA, the ADAS-Cog-13 has been extensively validated as an outcome measure in AD and MCI clinical trials, with established sensitivity to longitudinal change and disease progression (31,32). The convergence between the ADAS-Cog-13 cutoff identified in this epilepsy cohort and thresholds associated with early AD raises the possibility that elevated ADAS-Cog-13 scores in epilepsy may capture neurodegenerative vulnerability rather than epilepsy-related dysfunction alone. Rather than serving as a replacement for brief screening instruments such as the MoCA, the ADAS-Cog may be best positioned as a complementary tool for characterizing memory cognitive risk and informing longitudinal monitoring or clinical trial eligibility. Although the ADAS-Cog-13 requires greater administration time and examiner involvement than the MoCA, this added burden reflects its more granular assessment of memory and language, domains most relevant to cognitive aging and neurodegenerative risk in focal epilepsies.

An additional strength of the ADAS-Cog-13 is its extensive validation in AD research (33). In AD and MCI populations, ADAS-Cog-13 cutoff scores ranging from approximately 13 to 18 have been associated with early cognitive decline and progression risk, depending on cohort characteristics and analytic approach. The identification of an optimal cutoff of ≥15 in the present epilepsy cohort falls squarely within this range, raising the possibility that elevated ADAS-Cog-13 scores in epilepsy may reflect underlying neurodegenerative processes. A score above the cut-off may also indicate an increased risk of progression to AD. However, longitudinal studies examining progression to dementia in our epilepsy cohort are needed to determine the utility of this cutoff in predicting conversion.

Given accumulating evidence linking late-onset epilepsy and accelerated cognitive decline to increased AD risk, the ADAS-Cog-13 may offer a unique bridge between epilepsy and dementia research frameworks. Longitudinal studies incorporating biomarkers of Alzheimer’s pathology will be essential to determine whether elevated ADAS-Cog-13 scores in epilepsy predict subsequent neurodegenerative outcomes. Interestingly, ADAS-Cog-13 performance was largely independent of traditional epilepsy clinical variables, including age of onset, epilepsy duration, seizure activity, and number of ASMs, with the exception of a modest association with polytherapy. This finding aligns with prior work (22,34) suggesting that cognitive impairment in older adults with epilepsy is not solely explained by seizure burden or treatment factors, but instead reflects a multifactorial process (39).

Finally, we observed that male participants demonstrated more abnormal ADAS-Cog-13 performance than females. While exploratory, this finding is consistent with a prior systematic review reporting superior performance among women across several cognitive domains in epilepsy (40). Together, these observations warrant further investigation, as sex-specific cognitive trajectories in epilepsy remain poorly understood and may have implications for risk stratification and personalized screening approaches.

### Limitations and Future Directions

Several limitations should be acknowledged. First, the sample was predominantly White, highly educated, and composed of focal epilepsies of temporal and frontal onset, which may limit generalizability to more diverse populations and underscores the need for validation in cohorts with broader sociodemographic and epilepsy representation. Second, the cross-sectional design precludes conclusions regarding longitudinal cognitive change, trajectories of decline, or future dementia risk, key aspects of screening utility, particularly in clinical trial settings where detection of meaningful change over time is essential. Third, the modest sample size limited the ability to examine discrete IC-CoDE phenotypes with greater granularity, and larger cohorts will be required to more precisely evaluate screening performance across specific cognitive profiles. Fourth, although the ADAS-Cog-13 demonstrated reasonable diagnostic efficiency relative to IC-CoDE classifications, it should not be considered a substitute for a comprehensive neuropsychological assessment, rather it should be used to identify PwE that should be referred for neuropsychological evaluation. Finally, direct comparisons with other cognitive screening and composite measures, including more time-intensive instruments such as preclinical Alzheimer’s cognitive composites, were beyond the scope of this study but will be important as disease-modifying therapies emerge and indications for early detection expand.

### Conclusion

Given that progressive cognitive decline is a major concern in PwE, neuropsychological status is an outcome of major interest, and the availability of sensitive and specific cognitive metrics is critical for both clinical trials and clinical care. However, comprehensive neuropsychological testing is time-intensive, resource-dependent, and of variable accessibility globally. In epilepsy, there is no standard cognitive battery including among older adults (35), and, to our knowledge, the ADAS-Cog-13 has not been systematically evaluated as a screening instrument in older adults with epilepsy, nor benchmarked against an epilepsy-specific cognitive phenotyping framework.

From a clinical perspective, early identification of cognitive impairment is essential for optimizing treatment decisions, medication adherence, patient safety, financial management, and long-term planning in older adults(13–15) with epilepsy. This study demonstrates that the ADAS-Cog-13 is sensitive to cognitive impairment in this population when benchmarked against an epilepsy-specific gold standard, with particular strength in detecting memory-related deficits that are central to cognitive aging and neurodegenerative risk.

Beyond patient care, the ADAS-Cog-13 offers several advantages from a global health standpoint. Its extensive cross-cultural validation, availability in multiple languages, and widespread familiarity within international research infrastructure position it as a potentially scalable screening option in diverse epilepsy care settings, particularly where access to epilepsy-trained neuropsychologists is limited. While not a replacement for comprehensive neuropsychological assessment, the ADAS-Cog-13 may serve as a useful adjunct for identifying individuals at heightened cognitive risk and prioritizing referral and longitudinal monitoring.

PwE have been frequently excluded from AD clinical trials, hindering not only comparisons of PwE to ADRD cohorts on the ADAS-Cog-13, but also slowing understanding of the utility of AD treatments for PwE who develop AD. As disease-modifying therapies for ADRD emerge, tools capable of bridging epilepsy and dementia frameworks will become increasingly important. The ADAS-Cog-13 may represent one such bridge, supporting earlier detection, improved risk stratification, and more integrated care for older adults with epilepsy.

## Data availability statement

The authors have complete access to all study data and participant consent forms and assume full responsibility for the data, the research conduct, the analyses, the interpretation of the findings, and the right to publish all data. Data supporting the results of this study are available upon request from the study principal investigator (C.R.M.).

## IRB Statement

This study received approval from the Institutional Review Boards at the University of California, San Diego, Cleveland Clinic, and the University of Wisconsin-Madison. All participants provided written informed consent.

## Ethics approval statement

This study received ethical approval from the Institutional Review Boards at the University of California, San Diego, the Cleveland Clinic, and the University of Wisconsin-Madison.

## Patient consent statement

All participants provided written informed consent

## Author Contributions

JK, AR, BH conceptualized the idea. JK, AR, BH conducted the formal analyses. BH. JK, AR, IZ wrote the original draft. BPH, IZ, AR, RS, VP, WM, MW, RB, KA, LF, JJ, and CM supervised, validated, revised, reviewed, and edited the manuscript for intellectual content.

## Figure Legends

**Supplementary Table 1.** List and description of neuropsychological tests used for the BrACE study with normative means used in analysis and domain classification in IC-CoDE analysis.

| **Test Name** | **Description** | **Normative Means Used** | **IC-CoDE Domain Classification** |
| --- | --- | --- | --- |
| Alzheimer's Disease Assessment Scale – Cognitive Subscale (ADAS-Cog) Number Cancellation Task | Participants are given 45 seconds to cross off as many numbers as match two target numbers at the top of the page. The total score is the number of correct targets minus incorrect items minus the number of task reminders given. Total score is scaled 0-5, with 0 being the least amount of impairment. | ADNI-3 Healthy Control Sample | Attention/Processing Speed |
| Auditory Naming Test (ANT) | Participant responds as quickly as possible to 36 item descriptions with a correct target word. The score is the number of targets hit in under 20 seconds. | Hamberger et al. norms<sup>1</sup>, z-score | Language |
| Letter Fluency | Participant is given 60 seconds to say as many words as they can think of that start with certain letters of the alphabet (F, A, S). The total score is the number of words provided across all three trials. | Heaton norms, t-score | Executive Function |
| Multilingual Naming Test (MINT) | The participant tries to name 32 pictures of objects. The score is the total number of correct objects named. | UDS<sup>2</sup> Norms (Age, sex, and education), z-score | Language |
| Rey Auditory Verbal Learning Test, 30 Minute Delayed Recall | Participants learn 15 words across five immediate recall trials and a 30-minute delayed recall trial. The score is the number of words correctly identified for the delayed recall trial. | Meta-norms, z-score | Memory |
| Semantic Fluency | Participants are given 60 seconds to name as many animals as they can. The total score is the number of animals provided. | NACC<sup>3</sup> norms, z-score | Language |
| Trails Making Test Condition A | Participants are asked to connect the numbers 1-26 in ascending order on a sheet of paper. The score is the completion time in seconds. | NACC norms, z-score | Attention/Processing Speed |
| Trails Making Test Condition B | Participants are asked to switch between connecting numbers and letters in ascending order from I-M as quickly as possible. The score is the completion time in seconds. | NACC norms, z-score | Executive Function |
| Weschler Memory Scale (WMS) Logical Memory Story B Delayed Recall | Participants are read a short story and asked to recall as many details as possible both immediately and after a 20–30-minute delay. The score is the number of story elements recalled after the delay | NACC norms, z-score | Memory |
| Weschler Memory Scale (WMS) Visual Reproduction Delayed Recall | Participants are shown designs and are asked to draw those designs from memory immediately after the stimulus. The participants is instructed to redraw the designs from memory after a 20-30-minute delay. The score is the number of design elements correctly drawn after the delay. | Test Manual, t-score | Memory |
| 1. Hamberger, M. J., Heydari, N., Caccappolo, E., & Seidel, W. T. (2022). Naming in Older Adults: Complementary Auditory and Visual Assessment. *Journal of the International Neuropsychological Society*, 28(6), 574–587. https://doi.org/10.1017/S1355617721000552<br>2. UDS=Uniform Data Set<br>3. NACC=National Alzheimer's Coordinating Center |  |  |  |

**Supplementary Table 2.** Survey of Demographics Broken Down by IC-CoDE Classification and the statistical differences between Groups.

|  | IC-CoDE intact | IC-CoDE impaired | Statistic | p-Value |
| --- | --- | --- | --- | --- |
| **N** | 51 | 32 |  |  |
| **Years of Age** M (SD) | 66.08 (6.70) | 66.84 (7.14) | -0.494 | 0.311 |
| **Years of Education** M (SD) | 15.06 (2.65) | 14.59 (2.56) | 0.788 | 0.216 |
| **ADI National %** M (SD) | 47 (24.88) | 50.21 (28.88) | -0.520 | 0.302 |
| **Epilepsy Onset Age** (years, M (SD)) | 44.29 (19.39) | 41.52 (22.78) | 0.589 | 0.279 |
| **Epilepsy Classification: Late Onset (%)** | 15 (29.4) | 9 (29.0) |  | 0.588 |
| **Epilepsy Status: Active (%)** | 33 (66.0) | 20 (69.0) |  | 0.494 |
| **Epilepsy Duration** (years, M (SD)) | 21.78 (18.19) | 28.13 (23.09) | -1.318* | 0.097 |
| **Number of ASMs** M (SD) | **1.54 (0.86)** | **1.90 (0.96)** | **-1.733** | **0.044** |
| **Sex: Female (%)** | 31 (60.8) | 16 (50.0) |  | 0.230 |
| **Ethnicity: Hispanic (%)** | 3 (5.9) | 2 (6.3) |  | 0.644 |
| **Race: White (%)** | 47 (94.0) | 26 (83.9) |  | 0.138 |
| **Marital Status: Married (%)** | 35 (70.0) | 21 (67.7) |  | 0.558 |
| **ADAS-Cog-13 Total Score** M (SD) | **12.93 (4.95)** | **16.39 (4.62)** | **-3.172** | **0.001** |
| **Localization:** |  |  |  |  |
| **Frontal (%)** | 2 (3.9) | 2 (6.3) |  | 0.856 |
| **Temporal (%)** | 27 (52.9) | 14 (43.) |  |  |
| **Frontotemporal (%)** | 12 (23.5) | 9 (28.1) |  |  |
| **Unknown (%)** | 10 (19.6) | 7 (21.9) |  |  |

Note: \* indicates equal variances between groups were not assumed. Bolded statistics represent significant findings at a $p < .05$ .

**Supplementary Table 3.** Comparison of ADAS-Cog-13 Total Scores to Sociodemographic and Clinical Factors.

|  | **Statistic** | **p-Value** |
| --- | --- | --- |
| **Years of Education** | -2.14 | 0.052 |
| **Age** | -0.026 | 0.816 |
| **ADI National %** | 0.088 | 0.440 |
| **Age of Epilepsy Onset** | -0.025 | 0.820 |
| **Duration of Epilepsy** | 0.063 | 0.570 |
| **Number of ASMs** | 0.020 | 0.863 |
| **MoCA total Score** | **-0.403** | **<0.001** |
| **Sex** | **-1.986** | **0.025** |
| **Female** |  |  |
| **Male** |  |  |
| **Epilepsy Onset** | 0.125 | 0.450 |
| **Late Onset** |  |  |
| **Normal Onset** |  |  |
| **Epilepsy Status** | -0.554 | 0.291 |
| **Active** |  |  |
| **Controlled** |  |  |

Note: Bolded statistics represent significant findings at a $p < .05$ .

